# A Cascade of Care for Alcohol Use Disorder: Using 2015-2018 National Survey on Drug Use and Health Data to Identify Gaps in Care

**DOI:** 10.1101/2020.10.30.20222695

**Authors:** Carrie M. Mintz, Sarah M. Hartz, Sherri L. Fisher, Alex T. Ramsey, Elvin H. Geng, Richard A. Grucza, Laura J. Bierut

## Abstract

**Background:** Although effective treatments exist, alcohol use disorder (AUD) is undertreated. We used a cascade of care framework to understand gaps in care between diagnosis and treatment for persons with AUD.

**Methods:** Using 2015-2018 National Survey on Drug Use and Health data, we evaluated the following steps in the cascade of care: 1) prevalence of adults with AUD; 2) proportion of adults who utilized health care in the past 12 months; 3) were screened about alcohol use; 4) received a brief intervention about alcohol misuse; 5) received information about treatment for alcohol misuse; and 6) proportion of persons with AUD who received treatment. Analyses were stratified by AUD severity.

**Results:** Of the 171,766 persons included in the sample, weighted prevalence of AUD was 7.9% (95% CI 7.7-8.0%). Persons with AUD utilized health care settings at similar rates as those without AUD. Cascades of care showed the majority of individuals with AUD utilized health care and were screened about alcohol use, but the percent who received the subsequent steps of care decreased substantially. For those with severe AUD, 83.5% (CI: 78.3%-88.7%) utilized health care in the past 12 months, 73.5% (CI: 68.1%-78.9%) were screened for alcohol use, 22.7% (CI: 19.4%-26.0%) received a brief intervention, 12.4% (CI: 10%-14.7%) received information about treatment, and 20.5% (CI: 18%-23.1%) were treated for AUD. The greatest decrease in the care continuum occurred from screening to brief intervention and referral to treatment. More persons with severe AUD received treatment than were referred, indicating other pathways to treatment outside of the healthcare system.

**Conclusions:** Persons with AUD utilize health care at high rates and are frequently screened about alcohol use, but few receive treatment. Health care settings-particularly primary care settings-represent a prime opportunity to implement pharmacologic treatment for AUD to improve outcomes in this high-risk population.

## INTRODUCTION

Alcohol use disorder (AUD) is one of the most prevalent substance use disorders (SUDs) in the world and confers increased mortality risk. It is estimated that 93,000 people die of alcohol-related causes in the United States each year (Esser et al., 2020), and alcohol-related deaths are one of the leading causes of preventable death (Mokdad et al., 2004; U.S. Burden of Disease Collaborators, 2013). Of particular concern, mortality associated with AUD has increased in recent years (Case and Deaton, 2017; Spillane et al., 2020). Importantly, effective treatments for AUD exist. Evidence-based psychotherapy interventions, including motivational interviewing, cognitive behavioral therapy, and contingency management improve outcomes in persons with AUD (Carvalho et al., 2019; Knox et al., 2019), as does Alcoholics Anonymous (Kelly et al., 2020). In addition, there are three Food and Drug Administration (FDA)-approved medications for treatment of AUD–naltrexone (available in oral and extended-release depot formulations), acamprosate, and disulfiram. Naltrexone and acamprosate in particular have been shown to improve duration of sobriety and decrease heavy drinking days (Jonas et al., 2014), and thus pharmacotherapy is considered first-line treatment for persons with moderate or severe AUD (Kranzler and Soyka, 2018).

Health care providers are critical to the identification and treatment of persons with AUD. Since 1996, the U.S. Preventative Services Task Force (USPSTF) has recommended that physicians universally screen adult patients for alcohol misuse in health care settings and provide brief intervention for those with problem drinking (Bazzi and Saitz, 2018). These combined efforts–screening with brief intervention–are effective at decreasing self-reported problematic drinking among those with heavy drinking (Moyer et al., 2002; Willenbring, 2013). However, for those with AUD, screening and brief intervention has been shown to be insufficient (Willenbring, 2013), and referral to AUD treatment is recommended (Knox et al., 2019).

Encouragingly, there has been significant uptake of the USPSTF recommendation of screening for alcohol misuse. In a 2013 national survey, of those who met criteria for alcohol dependence and received care in ambulatory care settings, 81% reported being screened by a health care provider about alcohol use (Glass et al., 2016). However, far fewer reported receiving a brief intervention for problem drinking, with only 25% of respondents indicating that they were advised to cut down on their drinking or received information about treatment (Glass et al., 2016). Extending this work to 2013 and 2014 national data, Bandara et al. found that only 6.8% of those with AUD received any treatment for their alcohol use, which included mutual help such as Alcoholics Anonymous (Bandara et al., 2018).

To better understand the gaps in care between diagnosis and treatment, we can use an implementation science lens and apply a cascade of care framework. First developed in response to the HIV/AIDS epidemic (Gardner et al., 2011), a cascade of care tracks the proportion of the population of interest engaged in each step of a care continuum including diagnosis, engagement in medical care, receipt of treatment, retention in care and remission of disease (U.S. Department of Health & Human Services, 2020). A cascade of care framework has been adopted for other chronic diseases (Kazemian et al., 2019; Prabhakar and Kwo, 2019; Thomas, 2020), including opioid use disorder (Williams et al., 2019), to illustrate where gaps in care occur so that interventions can be targeted with the ultimate goal of decreasing mortality and improving disease outcomes.

Previous studies suggest there are significant gaps at multiple steps of an AUD cascade of care (Bandara et al., 2018; Glass et al., 2016; Hallgren et al., 2020). Critical to improving our understanding of a cascade of care for this population is increased knowledge of their patterns of health care utilization including ambulatory care visits, emergency room, and inpatient hospitalizations, so we can optimally focus intervention efforts. Health care utilization patterns may be especially pertinent for those with severe AUD, who are most likely to develop medical complications that require more acute levels of care.

Using data from the 2015-2018 National Survey on Drug Use and Health (NSDUH), we evaluated the following steps in the continuum of care, stratifying our analyses by AUD severity: 1) prevalence of U.S. adults with AUD in the general population; 2) proportion of adults who utilized health care in the past 12 months and 3) were screened about alcohol use by a health professional; 4) received a brief intervention about alcohol use from a health care professional; 5) received information about alcohol use and treatment for problems with alcohol use from a health care professional; and 6) proportion of adults with AUD in the general population who received treatment.

## MATERIALS AND METHODS

### Data Source

We analyzed NSDUH data from 2015-2018. NSDUH is conducted annually by the Substance Abuse and Mental Health Services Administration (Substance Abuse and Mental Health Services Administration, 2020) to measure substance use patterns in the United States; respondents are sampled from non-institutionalized, domiciled U.S. citizens aged 12 and older from all 50 states and the District of Columbia. The NSDUH provides nationally representative data on the prevalence and correlates of AUD and includes questions about health care utilization, alcohol screening, brief intervention, and referral to treatment as well as receipt of AUD treatment. Field workers interview participants in person; however, questions about potentially sensitive behaviors, including alcohol use, are administrated via audio-computer-assisted self-interview to maximize confidentiality. We focused our analyses on persons aged 18 years and older. Survey interview response rates ranged from 66.6% to 69.3% over the 2015-2018 period.

The study was exempted from human subjects review by the Institutional Review Board at Washington University School of Medicine.

### Cascade of Care Step 1: Prevalence of Mild, Moderate and Severe AUD

NSDUH uses DSM-IV criteria to generate diagnoses for past 12 month alcohol abuse and the more severe alcohol dependence. Under DSM-IV criteria, it is possible to endorse one symptom and meet criteria for alcohol abuse; a diagnosis of alcohol dependence requires endorsement of at least three of seven symptoms. DSM-5 criteria, which have been used clinically since 2013, removed the distinction between abuse and dependence and instead categorize AUD severity by the number of eleven possible symptoms endorsed to define mild, moderate and severe AUD. The NSDUH includes questions that map to ten of the eleven DSM-5 AUD criteria; respondents are not asked about craving for alcohol. Using similar methodology to that described by Johnson et al. 2020 (Johnson et al., 2020), we took a DSM-5-like graded severity approach, classifying persons with 0-1 symptom as not having AUD, those with 2-3 symptoms as having mild AUD, those with 4-5 symptoms as having moderate AUD, and those with 6 or more symptoms as having severe AUD. We used DSM-5 criteria to define past 12 month AUD severity for two reasons: DSM-5 is the current diagnostic system in use clinically, and by requiring endorsement of at least two symptoms to qualify for AUD, and six symptoms to qualify for severe AUD, we increased the likelihood of correctly classifying those with a clinically significant use disorder. See Supplementary Table 1 for the variables used to define AUD severity.

**Table 1.** Characteristics associated with past 12 month alcohol use disorder^±^ in the 2015-2018 National Survey of Drug Use and Health (NSDUH)

|  | **No AUD**<br>**(n=154,653)** | **Mild AUD**<br>**(n=11,408)** | **Moderate AUD**<br>**(n=3,356)** | **Severe AUD**<br>**(n=2,349)** |
| --- | --- | --- | --- | --- |
|  | **Weighted % (95% CI)** | **Weighted % (95% CI)** | **Weighted % (95% CI)** | **Weighted % (95% CI)** |
| **AUD Prevalence** | 92.1 (92.0-92.3) | 5.2 (5.1-5.3) | 1.6 (1.5-1.7) | 1.1 (1.0-1.1) |
| **Past 12 Month AUD Treatment** | 0.5 (0.5-0.6) | 2.6 (2.2-3.0) | 5.7 (4.4-6.9) | 20.5 (18.2-22.9) |
| **Sex** |  |  |  |  |
| Male | 47.1 (46.7-47.5) | 61.3 (59.8-62.7) | 63.2 (60.4-66.0) | 61.9 (59.1-64.6) |
| Female | 52.9 (52.5-53.3) | 38.7 (37.3-40.2) | 36.8 (34.0-39.6) | 38.1 (35.4-40.9) |
| **Age** |  |  |  |  |
| 18-25 years old | 13.1 (12.8-13.3) | 26.9 (25.9-27.9) | 23.2 (21.7-24.8) | 18.8 (17.2-20.4) |
| 26-34 years old | 15.3 (15.1-15.6) | 22.2 (21.0-23.4) | 23.9 (21.5-26.2) | 26.4 (23.9-28.8) |
| 35-49 years old | 24.7 (24.4-25.0) | 23.3 (22.0-24.6) | 27.4 (24.8-30.0) | 31.2 (28.2-34.2) |
| 50 or older | 46.9 (46.4-47.4) | 27.6 (25.9-29.3) | 25.5 (22.4-28.6) | 23.6 (20.7-26.5) |
| **Race/ethnicity** |  |  |  |  |
| Non-Hispanic White | 64.0 (63.5-64.5) | 64.4 (62.8-66.0) | 66.8 (64.2-69.4) | 64.7 (61.4-67.9) |
| Non-Hispanic Black | 11.9 (11.5-12.2) | 12.4 (11.6-13.2) | 9.2 (8.0-10.5) | 10.9 (9.2-12.6) |
| Non-Hispanic Other/Multi- Race | 8.2 (8.0-8.5) | 7.0 (6.2-7.9) | 7.1 (5.6-8.6) | 7.2 (6.0-8.5) |
| Hispanic | 15.9 (15.5-16.3) | 16.2 (14.9-17.5) | 16.9 (15.0-18.8) | 17.2 (14.1-20.3) |
| **Health Insurance** |  |  |  |  |
| Yes | 90.4 (90.2-90.7) | 87.4 (86.6-88.2) | 87.1 (85.6-88.6) | 83.5 (81.2-85.7) |
| **Past 12 Month Mental Health Treatment** | 13.7 (13.5-14.0) | 20.7 (19.5-21.9) | 27.3 (24.6-30.0) | 37.5 (34.7-40.2) |
| **Past 12 month illicit substance use disorder** | 2.0 (1.9-2.1) | 9.0 (8.3-9.8) | 17.0 (15.1-18.8) | 25.9 (23.8-27.9) |

Abbreviations: AUD= alcohol use disorder; CI=Confidence Interval.
<sup>‡</sup>AUD definition derived from *Diagnostic and Statistical Manual of Mental Disorders, 5<sup>th</sup> Edition* diagnostic criteria.

### Cascade of Care Step 2: Health Care Utilization

The NSDUH includes questions about past 12 month visits to ambulatory care settings, emergency room (ER) visits, and overnight inpatient hospitalizations; specific wording of questions is included in Supplementary Table 2. We recoded questions about utilization of each type of health care into dichotomous “yes”/”no” variables; we treated other responses as missing data. We then created a summary variable denoting whether a participant accessed at least one of these three health care settings in the past 12 months.

**Table 2.** Prevalence of past 12 month health care utilization by severity of alcohol use disorder^±^

|  | **No AUD**<br>**(n=154,653)** | **Mild AUD**<br>**(n=11,408)** | **Moderate AUD**<br>**(n=3,356)** | **Severe AUD**<br>**(n=2,349)** |
| --- | --- | --- | --- | --- |
|  | **Weighted % (95% CI)** | **Weighted % (95% CI)** | **Weighted % (95% CI)** | **Weighted % (95% CI)** |
| **Any Healthcare Utilization** | 84.2 (83.9-84.4) | 81.5 (80.6-82.5)<sup>***</sup> | 80.5 (78.4-82.7)<sup>***</sup> | 83.5 (81.3-85.7) |
| **Ambulatory Care Visit** | 82.6 (82.3-82.8) | 79.2 (78.2-80.2)<sup>***</sup> | 77.4 (75.1-79.6)<sup>***</sup> | 80.1 (77.9-82.3)<sup>*</sup> |
| **Emergency Room Visit** | 25.7 (25.3-26.1) | 28.3 27.2-29.4)<sup>***</sup> | 30.5 (28.2-32.8)<sup>***</sup> | 41.6 (38.1-45.1)<sup>***</sup> |
| **Overnight Hospitalization** | 10.1 (9.9-10.4) | 8.4 (7.6-9.2)<sup>***</sup> | 9.4 (7.9-10.9) | 16.6 (14.2-18.9)<sup>***</sup> |

Abbreviations: AUD= alcohol use disorder; CI=Confidence Interval.
<sup>±</sup>AUD definition derived from *Diagnostic and Statistical Manual of Mental Disorders, 5<sup>th</sup> Edition* diagnostic criteria.
<sup>\*</sup> $p<.05$ ; <sup>\*\*\*</sup> $p<.001$ . Reference group for each comparison was “No AUD” group.

### Cascade of Care Step 3: Alcohol Screening

Respondents who indicated accessing health care in the past 12 months were asked the following question: “During the past 12 months, did any doctor or other health care professional ask, either in person or on a form, if you drink alcohol?” Those who indicated that they had utilized health care and had consumed at least one alcoholic drink in the past 12 months were additionally asked to indicate whether the following happened in the past 12 months: “The doctor asked how much I drink;” “the doctor asked how often I drink;” and “the doctor asked if I have any problems because of my drinking.” We used similar methodology as Glass et al. (2016) and counted a positive response to any of these four items as indicative of having been screened for alcohol use. We intentionally used this broad definition to maximize sensitivity of our screening variable.

### Cascade of Care Step 4: Doctor or other health care professional provided brief intervention for drinking

Those who indicated that they had utilized health care and had consumed at least one alcoholic drink in the past 12 months were asked to indicate whether the following happened in the past 12 months: “The doctor advised me to cut down on my drinking.” Agreement with this statement was defined as receiving a brief intervention for drinking.

### Cascade of Care Step 5: Doctor or other health professional provided referral to treatment

Those who indicated that they had utilized health care and had consumed at least one alcoholic drink in the past 12 months were asked to indicate whether the following happened in the past 12 months: “The doctor offered to give me more information about alcohol use and treatment for problems with alcohol use.” Agreement with this statement was defined as receiving referral to treatment.

### Cascade of Care Step 6: AUD Treatment Receipt

Respondents who had consumed at least one alcoholic drink in the past 12 months were asked whether they had received treatment or counseling for use of alcohol or any drug, not including cigarettes, during the past 12 months. Those who answered yes were then asked whether the treatment was for alcohol use only, drug use only, or both alcohol and drug use. NSDUH used these responses to create a recoded dichotomous past 12 month alcohol treatment variable with the following responses: “yes” or “no/unknown.” We used this variable in our analyses (see Supplementary Table 2).

#### Analytic Plan

Analyses were conducted using SAS version 9.4 (Cary, NC, USA). To account for NSDUH’s complex sampling design, we applied survey weights created by NSDUH to all analyses so that estimates were representative of the national population. We used descriptive statistics to calculate observed proportions and corresponding confidence intervals for demographic variables, health care utilization patterns, and screening, brief intervention, and referral to treatment patterns. Chi Square statistics were used to examine differences in health care utilization by AUD severity.

#### Missing Data

Missing data were rare. There were 3,460 missing responses for past 12 month emergency room visits (2.0% of total sample) and 713 missing responses for past 12 month overnight hospitalizations (0.4% of total sample). There were 935 missing responses for the composite screening variable (0.5% of total sample) and 2,959 missing responses for the brief intervention and referral to treatment questions (1.7% of total sample). These missing responses were excluded from analyses.

## RESULTS

### AUD Prevalence and Sample Demographics

Demographic and clinical characteristics of the sample are presented in Table 1. Among the 171,766 persons included in the sample, the weighted prevalence of AUD as defined by DSM-5 criteria was 7.9% (95% CI 7.7-8.0%). Among those with AUD, mild AUD was most common and severe AUD was least common. As compared to those without AUD, persons with AUD were more likely to have a comorbid illicit drug use disorder and to have received mental health treatment in the past 12 months.

### Past 12 month Healthcare Utilization by AUD Severity

As shown in Table 2, the vast majority of respondents (>80%) reported utilizing at least one of the three healthcare settings queried, regardless of AUD diagnosis or severity. Those without AUD were statistically significantly more likely to report past 12 month healthcare utilization than those with mild or moderate AUD, however absolute prevalence differences were small. Importantly, there was no difference in overall healthcare utilization between those without AUD and those with severe AUD.

Ambulatory care settings were the most commonly accessed health care setting among all groups: more than 75% of persons in each group reported attending an outpatient appointment in the past year, with specific prevalence ranging from 77.4% (95% CI 75.1-79.6%) for persons with moderate AUD to 82.6% (95% CI 82.3-82.8%) for persons without AUD. Notably, persons with severe AUD were the most likely of all groups, and more than 60% likely than persons without AUD to have been hospitalized or gone to the ER in the past year.

### Screening, Brief Intervention, and Referral to Treatment (SBIRT) by AUD Severity

Table 3 shows the prevalence of alcohol screening, brief intervention, and referral to treatment among persons who reported utilizing health care in the past year. A majority of respondents reported being screened about alcohol use, and the likelihood of being screened increased with AUD severity, ranging from 75.3% (95% CI 74.9-75.8%) of those without AUD to 88.8% (95% CI 86.8-90.8%) of those with severe AUD. Brief intervention occurred much less frequently than screening, but also increased with AUD severity, ranging from 1.8% (95% CI 1.7-1.9%) of those without AUD to 27.7% (95% CI 24.2-31.1%) of those with severe AUD. Even fewer respondents with AUD were referred to treatment: 4.3% (95% CI 3.6-5.0%) of those with mild AUD, 6.8% (95% CI 5.5-8.2%) of those with moderate AUD and 15.1% (95% CI 12.4-17.7%) of those with severe AUD reported being given information by their healthcare provider about AUD treatment.

**Table 3.** Prevalence of alcohol screening, brief intervention, and referral to treatment by alcohol use disorder^±^ severity among those who utilized health care in the past 12 months.

|  | **No AUD**<br>**(n=123,395)** | **Mild AUD**<br>**(n=9,062)** | **Moderate AUD**<br>**(n=2,679)** | **Severe AUD**<br>**(n=1,921)** |
| --- | --- | --- | --- | --- |
|  | **Weighted % (95% CI)** | **Weighted % (95% CI)** | **Weighted % (95% CI)** | **Weighted % (95% CI)** |
| **Alcohol Screening** | 75.3 (74.9-75.8) | 85.1 (84.0-86.3) | 88.3 (86.6-89.9) | 88.8 (86.8-90.8) |
| **Brief Intervention** | 1.8 (1.7-1.9) | 10.2 (8.9-11.4) | 18.3 (16.4-20.1) | 27.7 (24.2-31.1) |
| **Referral to Treatment** | 0.9 (0.8-1.0) | 4.3 (3.6-5.0) | 6.8 (5.5-8.2) | 15.1 (12.4-17.7) |

Abbreviations: AUD= alcohol use disorder; CI=Confidence Interval.
<sup>‡</sup>AUD definition derived from *Diagnostic and Statistical Manual of Mental Disorders, 5<sup>th</sup> Edition* diagnostic criteria.

### Receipt of Treatment for AUD by AUD Severity

Overall, 5.7% of persons with AUD in our total sample reported receiving treatment. As shown in Table 1, prevalence of treatment increased with severity of illness: 2.6% (95% CI 2.2-3.0%) for mild AUD, 5.7% (95% CI 4.4-6.9%) for moderate AUD, and 20.5% (95% CI 18.2-22.9%) for severe AUD.

### Cascade of Care for AUD

Figure 1a-c shows the number of people who received each step in a cascade of care for severe, moderate and mild AUD. We present the weighted frequencies for each step to approximate trends on a population level. The number of individuals completing each step is relative to the baseline prevalence of AUD, not conditional on the number of individuals who completed the previous step.

**Figure 1a.**
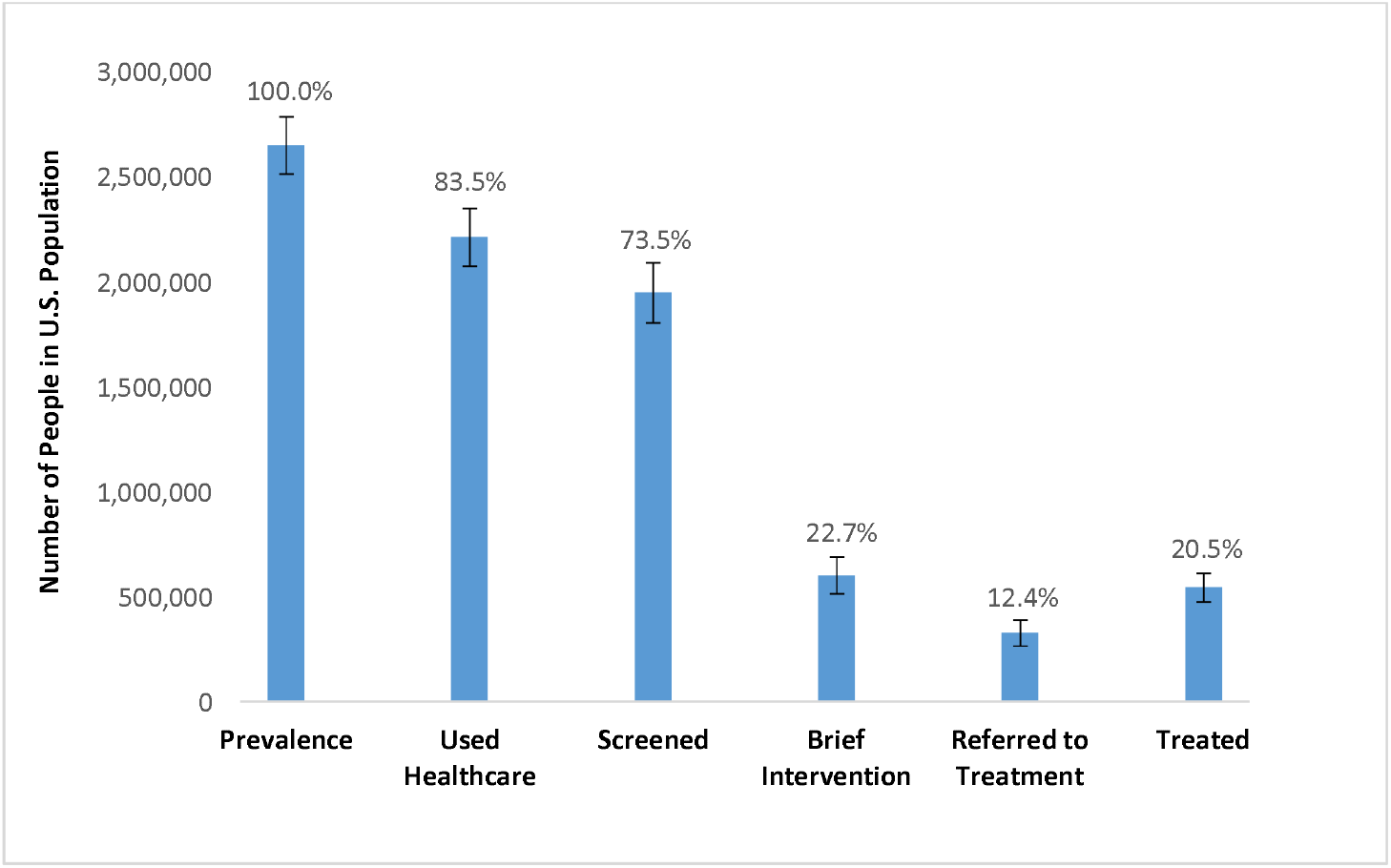
Cascade of care for severe alcohol use disorder: 2015-2018 National Survey of Drug Use and Health weighted prevalence data.

**Figure 1b.**
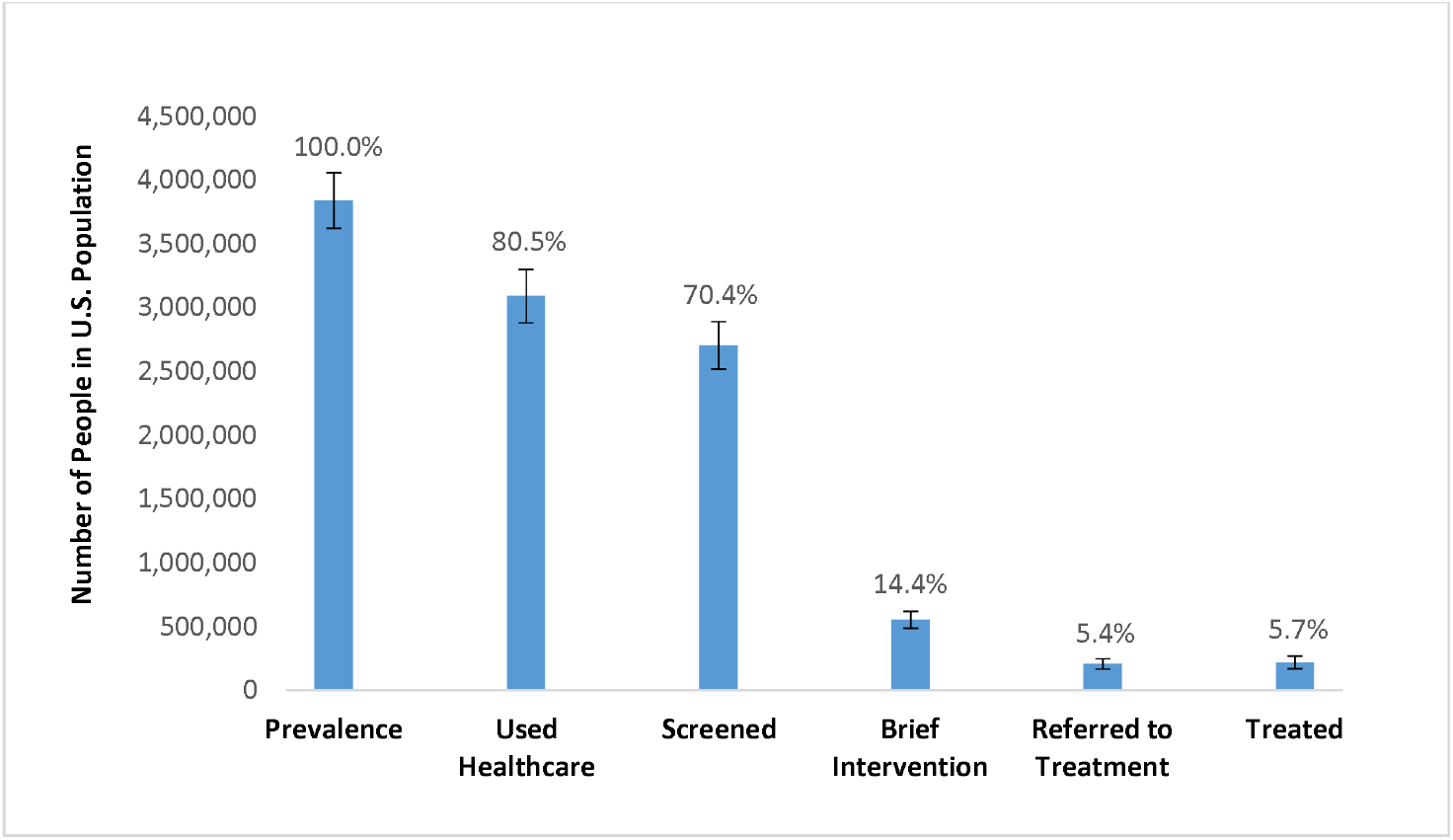
Cascade of care for moderate alcohol use disorder: 2015-2018 National Survey of Drug Use and Health weighted prevalence data.

**Figure 1c.**
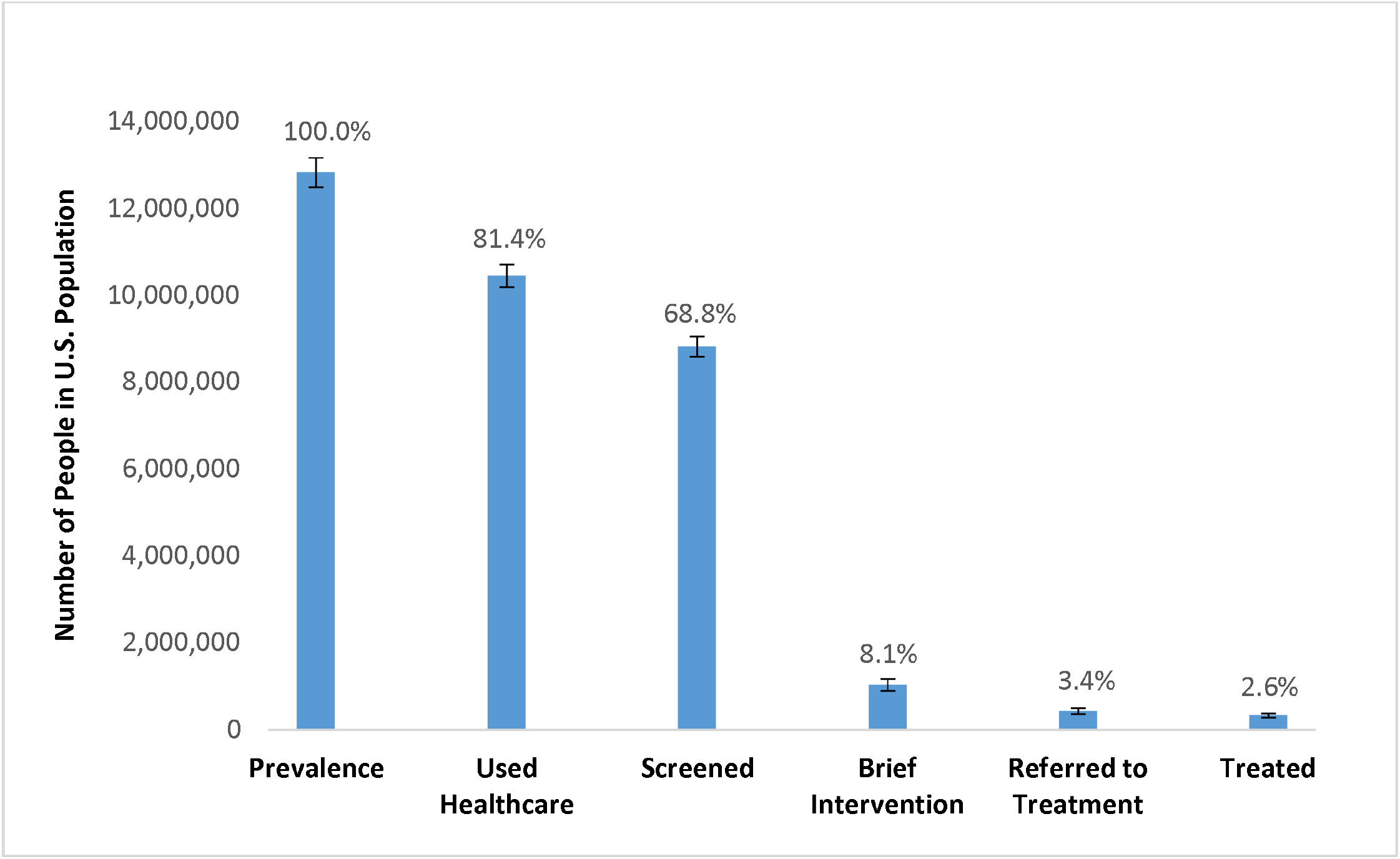
Cascade of care for mild alcohol use disorder: 2015-2018 National Survey of Drug Use and Health weighted prevalence data.

Similar patterns were observed within each AUD severity group: the majority of people utilized health care and were screened about alcohol use, but the proportion who received subsequent steps of care decreased substantially. Among the estimated 2,651,580 persons with severe AUD (Figure 1a.), 83.5% (CI: 78.3-88.7%, N=2,213,417) reported utilizing health care in the past 12 months. A majority of those with severe AUD (73.5%; CI: 68.1-78.9%, *N*=1,948,978) also reported being screened for alcohol use. An estimated 1,346,546 persons with severe AUD “fell off” the care continuum at the following step: only 22.7% (CI: 19.4-26.0%, *N*=602,432) reported being advised to cut down on drinking. An additional estimated 274,344 persons with severe AUD were lost from the care continuum following brief intervention: only 12.4% (CI: 10.0-14.7%, *N*=328,088) reported being referred to treatment. Conversely, there was an increase from referral to treatment to the last step in the cascade of care, with 20.5% (CI: 18.0-23.1%, *N*=544,497) reporting being treated for AUD in the past 12 months.

The cascades of care for those with moderate and mild AUD exhibited a similar pattern, though the number of individuals who fell off the care continuum from alcohol screening to the subsequent steps of brief intervention, referral to treatment, and receipt of treatment was even greater (Figures 1b and 1c, respectively).

## DISCUSSION

Individuals with AUD are woefully untreated. Using a cascade of care framework, we observed that this lack of treatment was not because individuals with AUD were unlinked to healthcare: Over 80% of individuals with AUD utilized health care in the past 12 months, and did so at similar rates as those without AUD. In addition, over 85% of persons with AUD who used health care reported being screened by a health professional about alcohol use. Thus, the high utilization of health care by individuals with AUD and the frequent screening for alcohol use during healthcare visits in the past 12 months presents the opportunity for widespread access to evidence-based AUD care.

However, despite the prevalence of alcohol screening, we found that the subsequent steps in the cascade of care were grossly underutilized, reiterating previous findings of large quality in care gaps for AUD (Bandara et al., 2018; Glass et al., 2016). Stratifying our analyses by severity of disease allowed for investigation of differing patterns of implementation gaps, however the overall cascade of care pattern was similar across severities. The gaps observed for severe AUD are particularly alarming and points to suboptimal care of a high-risk population. Those with severe AUD frequently have comorbid illnesses, and in our analysis they were the significantly more likely utilize the most expensive health care settings: emergency rooms and hospitals. Yet, even among this high-acuity group, only 20.5% reported receiving any AUD treatment in the past 12 months. The low prevalence of AUD treatment is especially striking when compared to treatment rates for other chronic diseases: indeed, recent cascade of care models for HIV and diabetes indicate that 64% of persons with HIV (HIV.gov, 2020) and 94% of persons with diabetes (Kazemian et al., 2019) receive treatment for their respective illnesses.

Our results add further support to the finding that pharmacologic treatment for severe AUD is underutilized given its documented efficacy (Jonas et al., 2014). Naltrexone and acamprosate have the best evidence for reducing alcohol consumption in AUD, as demonstrated by the number needed to treat (NNT), the average number of patients who need to be treated for one patient to receive a benefit. Naltrexone has been associated with an NNT of 20 for return to any drinking and an NNT of 12 for return to heavy drinking (Jonas et al., 2014). Acamprosate has been associated with an NNT of 12 for return to any drinking (Jonas et al., 2014). Yet, in an analysis of over 300,000 patients with AUD in the Veterans Health Administration healthcare setting from 2008-2009, fewer than 4% had documentation of pharmacologic treatment in the electronic medical record (Harris et al., 2012). More recently, Hallgren et al. (2020) showed that among persons with a documented diagnosis of AUD in a primary care setting, fewer than 10% received pharmacotherapy. Though we were not able to identify the prevalence of pharmacologic treatment for AUD in the current study (other than self-help groups, NSDUH does not contain questions about specific type of AUD treatment), only 20.5% of those with severe AUD reported any treatment, which provides a ceiling for the frequency of pharmacologic treatment in this criticially ill group. Given the known mortality associated with severe AUD (Laramee et al., 2015), this consistent low proportion of individuals with severe AUD who receive efficacious treatment, and particularly pharmacologic treatment, is disconcerting.

The implementation gap between healthcare screening for alcohol misuse and receipt of treatment highlights the fact that AUD treatment has historically been separated from mainstream health care settings and instead delivered via specialty care settings, including rehabilitation facilities, mental health centers, and non-health care settings such as peer support groups. Referral to specialized AUD treatment is an established approach in health care systems, and this approach is generally favorably viewed by health care providers who may prefer to refer patients to an expert rather than deliver AUD treatment themselves. Yet, this referral approach has significant drawbacks; chief among these is the low frequency with which individuals are successfully linked to treatment.

We therefore advocate for increased engagement of this high-risk population by implementing evidence-based treatment in the primary care setting. Primary care can be an effective setting for team-based AUD treatment at the-point-of-care, facilitated by electronic health records, and this approach has constituted a highly promising model for smoking cessation and opioid use disorder (Gunderson and Fiellin, 2008; Krantz and Mehler, 2004; National Institute on Drug Abuse, 2018; Ramsey et al., 2020; Ramsey et al., 2019a). This approach may involve a team of medical assistants and nurses conducting alcohol use assessment, providing brief advice about at risk alcohol use, and queuing an AUD medication order for the physician to prescribe. The physician can also reinforce further advice and encouragement and referral to further specialized treatment. Pharmacologic treatment for AUD in primary care is well-accepted by both patients and physicians (McNeely et al., 2018). The most commonly accessed health care setting reported by persons with AUD in our study was the ambulatory care setting, further underscoring its importance as a critical potential leverage point for providing AUD treatment (Anton et al., 2006; Grucza et al., 2020; Jonas et al., 2014; Knox et al., 2019; McNeely et al., 2018). Similar to other chronic illnesses, persons with severe AUD may also require specialized care, particularly for acute exacerbations; however, the need for specialty treatment should not preclude initiating pharmacologic treatment for many patients with AUD in mainstream health care settings.

Stigma associated with AUD from both patient and clinician perspectives has been cited multiple times as a barrier to treatment (Willenbring, 2013), and we argue that the separation of AUD treatment from mainstream health care settings has contributed to the this stigma. It is notable that approximately 25% of persons with severe AUD who received treatment indicated they had not utilized health care in the past year, suggesting individuals with severe AUD are finding their way to treatment through avenues other than the medical system. The reasons for this finding are likely multifactorial; however, the historic separation of SUD treatment from mainstream health care may be a contributing factor. In addition, physician time constraints and lack of knowledge about AUD treatment have also been cited as barriers (Rahm et al., 2015). As our group has observed for smoking cessation, there may also be a disconnect between physician and patient perspectives about the patient’s desire for care (Ramsey et al., 2019b). Whether this disconnect exists for AUD is an important area for future research.

As with all studies, there are limitations to this work. NSDUH does not sample those who are institutionalized, incarcerated, or homeless, and persons with AUD are overrepresented in those settings. Our findings regarding prevalence of screening and brief intervention efforts were given by self-report based on questions included in NSDUH. In addition, the NSDUH does not include detailed queries about type of treatment for AUD. Questions can also be raised about what constitutes treatment for AUD. For example, a brief intervention may be adequate treatment for those with mild AUD given the natural history of recovery in AUD (Dawson et al., 2005). Though these limitations exist, this large, general population sample nonetheless offers important insights into the care—and lack of care—for persons with AUD, and in particular for severe AUD.

## Conclusions

Persons with AUD utilize health care at high rates that are similar to those without AUD. Most persons with AUD report being screened about alcohol use, however only a minority receive AUD treatment.

Ambulatory care settings-the most commonly used form of health care for persons with AUD-represent a prime opportunity to implement point-of-care pharmacologic treatment for AUD to improve outcomes in this high-risk population.

## Data Availability

Data were from the 2015-2018 National Surveys on Drug Use and Health, which are publically available.

https://www.datafiles.samhsa.gov/study/national-survey-drug-use-and-health-nsduh-2002-2018-nid18771

## ACKNOWLEDGMENTS

We thank Jingling Chen for her assistance creating figures.

