## Supplementary Tables for "A Cascade of Care for Alcohol Use Disorder: Using 2015-2018 National Survey on Drug Use and Health Data to Identify Gaps in Care"

Supplementary Table 1: NSDUH variables used to define DSM-5 alcohol use disorder (AUD)

| **DSM-5 Criteria** | **Variable** | **Question** | **Survey Question Text** |
| --- | --- | --- | --- |
| 1: Alcohol is often taken in larger amounts or over a longer period than was intended. | ALCKPLMT | DRALC05 | DRAL04: During the past 12 months, did you try to set limits on how often or how much alcohol you would drink?  DRALC05 asked if DRALC04=Yes  Were you able to keep to the limits you set, or did you often drink more than you intended to?  1=Usually kept to limits set  *2=Often drank more than intended |
| 2: There is a persistent desire or unsuccessful efforts to cut down or control alcohol use. | ALCCUTEV | DRALC09 | DRAL08: During the past 12 months, did you want to or try to cut down or stop drinking alcohol?  DRALC09 asked if DRALC08=Yes  During the past 12 months, were you able to cut down or stop drinking alcohol every time you wanted to or tried to?  1=Yes  *2=No |
| 3: A great deal of time is spent in activities necessary to obtain alcohol, use alcohol, or recover from its effects. | ALCLOTTM | DRALC01 | During the past 12 months, was there a month or more when you spent a lot of your time getting or drinking alcohol?  *1=Yes  2=No |
|  | **OR** | | |
|  | ALCGTOVR | DRALC02 | During the past 12 months, was there a month or more when you spent a lot of time getting over the effects of the alcohol you drank?  *1=Yes  2=No |
| 4: Craving, or a strong desire or urge to use alcohol. | N/A | N/A | NOT ASSESSED |

| **DSM-5 Criteria** | **Variable** | **Question** | **Survey Question Text** |
| --- | --- | --- | --- |
| 5: Recurrent alcohol use resulting in a failure to fulfill major role obligations at work, school, or home. | ALCSERPB | DRALC18 | Sometimes people who drink alcohol have serious problems at home, work or school — such as:   - Neglecting their children - Missing work or school - Doing a poor job at work or school - Losing a job or dropping out of school   During the past 12 months, did drinking alcohol cause you to have serious problems like this either at home, work, or school?  *1=Yes  2=No |
| 6: Continued alcohol use despite having persistent or recurrent social or interpersonal problems caused or exacerbated by the effects of alcohol. | ALCFMCTD | DRALC22 | Did you continue to drink alcohol even though you thought your drinking caused problems with family or friends?  *1=Yes  2=No |
| 7: Important social, occupational, or recreational activities are given up or reduced because of alcohol use. | ALCLSACT | DRALC17 | This question is about important activities such as working, going to school, taking care of children, doing fun things such as hobbies and sports, and spending time with friends and family.  During the past 12 months, did drinking alcohol cause you to give up or spend less time doing these types of important activities?  *1=Yes  2=No |
| 8: Recurrent alcohol use in situations in which it is physically hazardous. | ALCPDANG | DRALC19 | During the past 12 months, did you regularly drink alcohol and then do something where being drunk might have put you in physical danger?  *1=Yes  2=No |
| 9: Alcohol use is continued despite knowledge of having a persistent or recurrent physical or psychological problem that is likely to have been caused or exacerbated by alcohol. | ALCEMCTD | DRALC14 | Did you continue to drink alcohol even though you thought drinking was causing you to have problems with your emotions, nerves, or mental health?  *1=Yes  2=No |
|  | **OR** | | |
|  | ALCPHCTD | DRALC16 | DRALC15: During the past 12 months, did you have any physical health problems that were probably caused or made worse by drinking alcohol?  DRALC16 asked if DRALC15=Yes  Did you continue to drink alcohol even though you thought drinking was causing you to have physical problems?  *1=Yes  2=No |

| **DSM-5 Criteria** | **Variable** | **Question** | **Survey Question Text** |
| --- | --- | --- | --- |
| 10: Tolerance, as defined by either of the following: a) A need for markedly increased amounts of alcohol to achieve intoxication or desired effect; b) A markedly diminished effect with continued use of the same amount of alcohol. | ALCNDMOR | DRALC06 | During the past 12 months, did you need to drink more alcohol than you used to in order to get the effect you wanted?  *1=Yes  2=No |
|  | **OR** | | |
|  | ALCLSEFX | DRALC07 | During the past 12 months, did you notice that drinking the same amount of alcohol had less effect on you than it used to?  *1=Yes  2=No |
| 11: Withdrawal, as manifested by either of the following: a) The characteristic withdrawal syndrome for alcohol; b) Alcohol (or a closely related substance, such as a benzodiazepine) is taken to relieve or avoid withdrawal symptoms. | ALCWD2SX | DRALC11 | Please look at the symptoms listed below. During the past 12 months, did you have 2 or more of these symptoms after you cut back or stopped drinking alcohol?   - Sweating or feeling that your heart was beating fast - Having your hands tremble - Having trouble sleeping - Vomiting or feeling nauseous - Seeing, hearing, or feeling things that weren’t really there - Feeling like you couldn’t sit still - Feeling anxious - Having seizures or fits   *1=Yes  2=No |

Positive responses to each criterion (denoted by * in the table above) were added to arrive at a score of 0 to 10 (craving was not assessed). All missing data were coded to NO.

0-1 = No AUD

2-3 = Mild AUD

4-5 = Moderate AUD

6 or more = Severe AUD

Supplementary Table 2: NSDUH variables used for analysis

| **Variable** | **Question** | **Survey Question Text** |
| --- | --- | --- |
| NMERTMT2 | HLTH16 | During the past 12 months, that is since DATEFILL, how many different times have you been treated in an emergency room for any reason? |
| INHOSPYR | HLTH17 | During the past 12 months, have you stayed overnight or longer as an inpatient in a hospital? |
| NMVSOPT2 | HLTH19 | During the past 12 months, how many times have you visited a doctor, nurse, physician assistant or nurse practitioner about your **own** health at a doctor’s office, a clinic, or some other place? |
| HPUSEALC | HLTH20b | During the past 12 months, did any doctor or other health care professional ask, **either in person or on a form**, if you drink alcohol? |
|  | HLTH22 | Please think about all of the talks you have had with a doctor or other health care professional during the past 12 months when you answer this question. Choose the statement or statements below that describe any discussions you may have had **in person** with a doctor or other health professional about your **alcohol use**.  To select more than one statement, press the space bar between each number you type. When you have finished, press [ENTER]. |
| HPALCAMT | HLTH221 | The doctor asked how much I drink. |
| HPALCFRQ | HLTH222 | The doctor asked how often I drink |
| HPALCPRB | HLTH223 | The doctor asked if I have any problems because of my drinking. |
| HPALCCUT | HLTH224 | The doctor advised me to cut down on my drinking. |
| HPALCTX | HLTH225 | The doctor offered to give me more information about alcohol use and treatment for problems with alcohol use. |
| HPALCNOT | HLTH226 | The doctor didn’t discuss my alcohol use with me in the past 12 months. |
| TXYRALC | Derived from TX02  (variable name TXYRRECVD)  Derived from TX03  (variable name TXYRALDGB) | During the past 12 months, that is, since [DATEFILL], have you received treatment or counseling for your use of alcohol or any drug, not counting cigarettes?”  IF YES TO TX02, RESPONDENT IS ASKED TX03:  During the past 12 months when you received treatment, was the treatment for alcohol use only, drug use only, or both alcohol and drug use? |
